# Should COVID-specific arrangements for abortion continue? The views of women experiencing abortion in Britain during the pandemic

**DOI:** 10.1101/2022.02.17.22271080

**Authors:** Patricia A. Lohr, Maria Lewandowska, Rebecca Meiksin, Rachel Scott, Jennifer Reiter, Natasha Salaria, Sharon Cameron, Melissa Palmer, Rebecca French, Kaye Wellings

## Abstract

**Background:** During the COVID-19 pandemic, the British governments issued temporary approvals enabling the use of both pills for medical abortion at home. This permitted the introduction of a fully telemedical model of abortion care with consultations taking place via phone or video call and medications delivered to women’s homes. The approvals in England and Wales will expire at the end of March 2022, while that in Scotland remains under consultation.

**Methods:** We interviewed 30 women who had undergone an abortion in England, Scotland or Wales between August and December 2021. We explored their views on the changes in abortion service configuration during the pandemic and whether abortion via telemedicine and use of abortion medications at home should continue.

**Results:** Support for continuation of the permission to use mifepristone and misoprostol at home was overwhelmingly positive. Reasons cited included convenience, comfort, reduced stigma, privacy, and respect for autonomy. A telemedical model was also highly regarded for similar reasons but for some its necessity was linked to safety measures during the pandemic and an option to have an in-person interaction with a health professional at some point in the care pathway was endorsed.

**Conclusions:** The approval to use abortion pills at home via telemedicine are supported by women having abortions in Great Britain. The respective governments in England, Scotland, and Wales, should be responsive to the patient voice and move to make permanent these important advances in abortion care.

**What is already known on this topic:** During the COVID-pandemic, specific permission to use both pills for medical abortion at home was granted in England, Scotland and Wales leading to the widespread implementation of a telemedical model with direct-to-patient delivery of medications. The safety, effectiveness, and acceptability of this model of care had been well-documented prior to and during the pandemic.

**What this study adds:** This study adds the voices of women undergoing abortion during the pandemic regarding the specific changes that led to the transformation of medical abortion care in Britain. Amongst 30 women interviewed, there was endorsement for the continuation of permissions to use medical abortion pills at home via telemedicine.

**How this study might affect research, practice, or policy:** The UK government’s vision of health provision puts patients and the public first, where “no decision about me, without me” is the norm. Our findings support law and policy makers in applying this principle to recent developments in abortion care by making the permissions permanent.

## INTRODUCTION

Significant changes to the nature and context of abortion provision are taking place in the UK, many of which were accelerated by the COVID pandemic. Following the licensing of the anti-progestogen mifepristone in 1991, the proportion of abortions performed using medication instead of surgery has risen year on year in Britain.(1) The most used regimen involves a single oral dose of mifepristone followed 24-48 hours later by sublingual, buccal, or vaginal administration of misoprostol, which induces pregnancy loss in a process very similar to miscarriage.(2) Prior to the pandemic, for pregnancies under 10 weeks’ gestation, women could use misoprostol at home but were required by law to attend a registered abortion clinic or NHS hospital to take mifepristone.(3)

To reduce the risk of COVID-19 transmission and ensure continued access to abortion services, in March 2020 temporary approvals were granted by the governments in England, Scotland and Wales to permit both mifepristone and misoprostol to be used at home.(4–6) Mifepristone could be used at home to 9 weeks and 6 days gestation in England and Wales and 11 weeks and 6 days gestation, as determined by guidance, in Scotland.(7) Alongside, the Royal College of Obstetricians and Gynaecologists released COVID-specific guidance for abortion services to implement a telemedical service delivery model consisting of consultations by phone or video call, estimation of gestational age using last menstrual period instead of routine ultrasound, and direct-to-patient delivery of abortion pills.(8)

Following the approvals, use of medical abortion increased, accounting for 88% of abortions in England and Wales from April-December 2020 compared to 77% in January-March 2020.(1) Forty-seven percent of early medical abortions were carried out with home-use of both medications. An associated decline in the gestational age at abortion was observed, attributed to faster access to treatment with the telemedical model.(9)

The dispensation to allow home administration of both abortion medications was planned to end when the provisions of the Coronavirus Act 2020 expire, or two years after the decision was made, whichever was sooner. At the time of writing, discussions are in progress to determine whether the guidance should remain in place after this time. Crucial to those deliberations are the perceptions of women themselves. In this paper, we report on the views of 30 women in Britain on whether the COVID-related measures should continue, drawing on their experience of abortion during a period in which it applied.

## METHODS

### Sampling

A purposive sample of 30 women with recent (past 2-8 weeks) experience of abortion were recruited between July and December 2021 from six sites: three independent-sector services commissioned by the NHS (a telemedicine hub in the North of England and in-person clinics in London and South West England); and NHS sites in Scotland, Wales and England (London). Inclusion criteria were: age 16 and over, able to give informed consent, UK-resident, and choosing abortion for reasons other than fetal anomaly.

Clinic or telehub staff introduced the study to potential participants after consent for the abortion was obtained and, with permission, passed the details of those interested in participating to team researchers for follow up, when willingness to take part was ascertained and a time for the interview established. Women having hospital-based abortions were recruited at initial assessment through their provider.

### Data collection

Semi-structured, in-depth interviews were carried out by phone or using video-conference software according to participant’s preference and, with their permission, audio-recorded and transcribed. Consent was recorded in the interview. A £20 high street voucher was offered in appreciation for their time. The interview guide captured women’s accounts of their recent experience of abortion. Opinions were specifically sought on whether temporary COVID-related regulations governing provision should be permanent; the reasons for their views and how they related to their own experience.

### Data analysis

Data were analysed the Framework Method.(10) An initial matrix was created into which summary data were entered, by case and by code. Codes of specific relevance to the study aims and research questions were applied *a priori* to the entire data set. Transcripts were read and coded by pairs of researchers (ML, PL, RM, JR, RS, KW) and interpretive themes emerging inductively from the data were then identified, shared, and agreed and added iteratively as analysis progressed, going back and forth between data and interpretation.

### Ethical approval

Ethical approvals were obtained from the Research Ethics Committees of the British Pregnancy Advisory Service (reference 2021/02/WEL), the London School of Hygiene & Tropical Medicine (reference 22761) and the NHS (reference 21/LO/0236).

### Patient and Public Involvement

The central aim of this research was to allow patient voices to guide the design of services. A PPI panel, established for the study, helped inform study design and commented on the interview guide and Participant Information Sheet.

## RESULTS

We interviewed 30 women aged 21 to 43 years old. Four had had surgical abortion and 26 a medical abortion. The characteristics of the participants are presented in Table 1.

**Table 1.** Characteristics of the sample.

| **ID** | **Abortion method** | **Age** | **Ethnicity** | **Parity** | **Previous abortions** | **Country** |
| --- | --- | --- | --- | --- | --- | --- |
| **01** | Medical at home | 31-35 | White British | No | No | England |
| **02** | Medical at home | 26-30 | White British | No | No | England |
| **03** | Medical at home | 21-25 | British Bangladeshi | No | Yes | England |
| **04** | Medical at home | 26-30 | White British | No | No | England |
| **05** | Medical at home | 36-40 | White British | Yes | Yes | England |
| **06** | Medical at home | 36-40 | White British | Yes | Yes | England |
| **07** | Medical at home | 41-45 | White British | Yes | Yes | England |
| **08** | Medical at home | 31-35 | White British | No | Yes | England |
| **09** | Medical at home | 21-25 | White British | No | No | Wales |
| **10** | Medical at home | 31-35 | White British | No | Yes | Scotland |
| **11** | Medical at home | 21-25 | White British | No | No | Scotland |
| **12** | Medical at home | 26-30 | White Irish | No | Yes | England |
| **13** | Medical at home | 21-25 | White British | No | No | England |
| **14** | Medical at home | 26-30 | White British | Yes | No | Scotland |
| **15** | Medical at home | 31-35 | White British | Yes | No | Scotland |
| **16** | Medical at home | 31-35 | White Polish | No | No | Scotland |
| **17** | Medical at home | 26-30 | White British | No | No | Scotland |
| **18** | Medical at home | 36-40 | White Canadian | No | No | Scotland |
| **19** | Medical at home | 21-25 | White British | No | No | Scotland |
| **20** | Surgical | 36-40 | Not specified, Hungarian | Yes | Yes | England |
| **21** | Medical at home | 36-40 | Not specified | Yes | No | Scotland |
| **22** | Medical at home | 26-30 | White British | No | Yes | Scotland |
| **23** | Surgical | 26-30 | White British | No | Yes | Scotland |
| **24** | Medical at home | 26-30 | White British | No | No | Scotland |
| **25** | Medical at home | 26-30 | White British | Yes | Yes | England |
| **26** | Surgical | 21-25 | White British | No | No | England |
| **27** | Surgical | 31-35 | White British | Yes | No | Scotland |
| **28** | Medical at home | 26-30 | White Hungarian | No | Yes | Scotland |
| **29** | Medical at home | 21-25 | Pacific Islander | No | No | Scotland |
| **30** | Medical at home | 31-35 | White British | No | No | England |

### Awareness of COVID-related arrangements and views on their continuation

Not all women had been aware of the impact of COVID on abortion provision, but support for the continuation of the revised delivery model was near universal. Only one woman in the sample demurred and she was against medical abortion in any form and had herself had a surgical abortion. Two women were unsure. Supportive responses included: *“I think 100% they should continue with this”* [05]; *“Without a doubt, this is the way forward”‘* [06]; *“I can’t see any benefit to going back to the old way of doing things”* [07]; *“I really strongly feel it shouldn’t change”* [22]; and *“Definitely being able to do it on my phone and by post. You can’t compare*.*”* [30]. Women considered themselves fortunate to have been offered home management of both medications and saw it as a fortuitous, if unintended, consequence of the pandemic.

### Factors influencing opinion

#### Convenience

Foremost among reasons for favouring continuation of the current model of care was convenience.

> *The fact that I could do it … at my convenience, and …because I just had to pop in and pick up a package… I think it’s brilliant. I think it’s amazing*.” [10]

Being able to plan the timing of the treatment around work and childcare was a major advantage.

> *It meant that I didn’t have to take any days off work. I didn’t have to tell work anything, and that suited me, because I didn’t want anyone to know … it worked really well, and I was able to fit it into my life, rather than go out of my way and make it a bigger deal than what I wanted it to be*. [02]
>
> *I think there should be the option of taking it at home. For a lot of women, it’s hard to balance their work life. In addition to stress that it can cause to your general life, and physically go to the clinic that may not be very close to you…especially if you’ve got other responsibilities at home such as children or caring duties, that option is very workable around your life*. [03, comparing her recent abortion with a previous one)

Abortions were often scheduled to take place at the weekend, obviating the need for time off work and disclosure to colleagues, also ensuring availability of support from friends and partners or - for those living with parents or family - a time when privacy was assured - advantages recognised by clinic staff as well as patients.

> *They told me on the phone… “If you take the first pill on Thursday, and then do the second ones on Saturday then that means you can do it at the weekend’*. [10]

Benefits were perceived for saving and planning time. Women with previous experience of abortion compared time wasted in travel, in repeat appointments and in lengthy waits at clinical facilities with the efficiency of the current arrangements.

> *If it was something that could remain, I would definitely prefer it…because you’ve got to go to so many appointments… you’ve got your original consultation and then you have to go back again for the first pill, and then for the second one, you would go back the next day*. [12]
>
> *Where I live, it’s a good hour away from where the clinic is. So, with two kids the first time around, it was quite hard to get there because I’m sure it was two days in a row, not everything happened in the first day. The second time around [recalling a previous abortion] was a lot easier because everything was over the phone, through email, and it got posted out within the week after the final phone call*. [25]

The new arrangements were also seen to as cost-saving, for women themselves through savings in travel costs and for the health service: “*It’s probably much cheaper and more effective to just post it out*”. [11]

Women commonly acknowledged that the benefits of the COVID-related arrangements would be even greater for those less advantaged, who might lack flexibility at work, opportunities for childcare, or the means to travel.

#### Home comforts

Home management was preferred by many for the comfort and privacy afforded: “*I preferred to be in my own space*” [12]; *“… you want to lie in bed with a hot water bottle and just be on your own…”* [15]; “*If you’re going to be upset, do not have an audience*” [06], “*it was great that I could take the four pills in my own home and in my own bed*” [16]. Home management was also seen to reduce the possibility of encountering judgmental attitudes, whether on the part of clinical staff, people they knew, or protesters.

> *Even though that’s people’s jobs, you don’t know how they feel personally about it…. So there’s that fear of being judged, of feeling ashamed. To not have to actually see anyone face to face, was really great actually*. [14]
>
> *There’s people that are against abortions…What if it’s in your local area and people see you go in there?* [04]

#### Increased autonomy

The self-regulation afforded by home management was also welcomed. Typical comments included: “*you’re in control of the situation*” [26]; “*it’s on your terms*” [06]; “*it just felt like it was much more like my decision … wasn’t being done to me or anything like that*”. [18] “*It was completely up to me, it was very much my choice, my body […] I took care of my own needs*.” [07] Autonomy was considered especially important to women who had thus far seen themselves as lacking agency in their lives. “*It’s not a great situation but it made it better by being able to have complete control of my own thing*”. [07]

Such views were widely expressed in support of continuation of COVID-related arrangements, but were tempered by comments from some to the effect that there was a balance to be struck between user autonomy and provider support. Whilst many women felt relief and satisfaction at being able to self-manage their abortion, others would have welcomed a watchful eye. Their comments suggested that a little more oversight would have been welcome at various points in the care pathway, for example, to allay doubts over the ‘rightness’ of their decision; reassurance during the abortion that what was experienced was normal; and pro-active call backs after the procedure to check that everything was fine. Some women had been able to collect the medication from clinics instead of having them posted, an option welcomed as allowing “*at least one point of face-to-face contact with someone*.” [05]

### The importance of choice

A strong consensus emerged on the need for choice, expressed in comments such as “*I think you should have the option if you want* “[19]; “*I guess it’s just down to the individual person’s comfort*” [04]; “*I think the best thing you can do is give people a choice […] give as many different options as possible*.” [13] The importance of tailoring care to individual needs was stressed.

> *I can see the benefits of both […] I might have preferred to actually take the tablet in the clinic because it feels almost like I’ve got hands to hold. […] But there’s some women who might […] want to take that time and do it when they feel ready. […] It’s not a one size fits all approach, everyone is in different circumstances, […]. To have the flexibility and the option for the woman to choose the way they prefer to do actually might be more beneficial*. [09]

While virtually all the women interviewed, even those with less favourable experiences, saw the benefits of the COVID arrangements as outweighing the costs, there was no sense that they should entirely replace previous models of care. Even those whose needs were fully met by remote support and home administration were able to envisage circumstances in which this might not be so. For women unable to be open about their abortion; for those less able to follow the instructions; and for those with less agency; the option of facility-based care alongside self-administration was seen to be optimal.

> T*he option to go to the clinic should still be available because if you have a husband who opens your post then that can cause problems* [07]
>
> *For those that maybe have anxiety about not taking the pill at the right time, [or the] right way, being supervised while doing it works for them. So I think having that choice is really good*. [03]

The dominant view then was that the current guidelines should be continued, not as a replacement but as an alternative option, to what went before.

> *I personally think that having options is great. So if someone wants to do it at home, then they should be allowed to. If they want do it in the clinic, they should be allowed to do that as well. Because everyone, everyone’s different. I think giving people the choice is probably the […] ideal situation*. [29]

## DISCUSSION

### Summary of findings

Continuation of the current COVID-related measures for abortion provision beyond March 2022 was emphatically and near-unanimously endorsed by women taking part in this study. Receipt of both abortion medications by post or collection, followed by home administration and management, was seen to save time and money, afford greater privacy and comfort, grant women more autonomy, and to be more easily accommodated into everyday life. Any shortcomings of the current arrangements, in terms of unmet need for in-person care and support, were seen as remediable through hybrid systems of abortion care, combining elements of home and facility-based care; self-administration and provider assistance; and in-person and virtual support. The concept of choice was considered of utmost important as was the need to tailor models of abortion care to individual circumstances and preferences.

### Strengths and limitations

Our study is unique in being the only Britain-wide study examining women’s views on the current COVID-related arrangements and policies for abortion with participants drawn from both independent providers of abortion care and NHS facilities. The methodological approach taken allowed women’s voices to be heard directly, providing access to their priorities and preferences, and so enhancing understanding of the meaning and significance of abortion experience for them.

A weakness of this analysis is that it did not include women aged under 20, thereby missing the opportunity to understand specific challenges or benefits in this age group. Further, women volunteering for interview may have occupied more extreme ends of the satisfaction spectrum, introducing a bias towards more positive or more negative views and experiences.

### Contextualisation and interpretation

The COVID pandemic has been an impediment to sexual and reproductive health services but it has also provided the impetus to new ways of working, an effect described as the ‘COVID-19 silver lining’.(11) In the context of abortion provision it has served to provide what might be seen as a ‘natural experiment’ in which the merits of a new model of care can be observed in the general population. Evaluation of the use of mifepristone as well as misoprostol at home pre-dates the COVID pandemic and is based on earlier experimental and quasi-experimental studies, and those based on selective samples such as women obtaining abortion medication online. The findings of these studies, that home management is feasible and acceptable,(12–15) are consistent with those of our ‘real world’ study.

Our findings echo those of previous studies during the pandemic period demonstrating that self-management of early medical abortion at home, supported by telemedicine, is acceptable to most women who received this model of care.(16–18) No other studies, to our knowledge, have specifically asked women for their opinion on whether the current arrangements should continue after the period specified, but quantitative research based on samples recruited from specific abortion providers indicate that this would be the future method of choice for a high proportion of women.(17,18) The view captured in our study that there should continue to be a choice of model of care is reflected in other research; one in five patients indicating a preference for inclusion of some face-to-face interaction.(18)

### Implications for policy and practice

This study has shown self-managed early medical abortion facilitated by telemedicine to be not only acceptable to, but preferred by, many women. Other research has also shown it to be safe (19) and cost-effective, resulting in significant cost savings for the NHS.(20) This model of care of aligns with broader trends within 21st century health systems: recognition of the need for patient-centred approaches, shared decision making in health care, and supported self-management, principles supported by advances in digital technology.(21)

The strong plea from women in our study for choice must be heeded. Not all women are eligible for medical abortion and self-management is not feasible for all. The evidence from this study is that choice is important to women and needs to be interpreted not only in terms of alternatives to home management, but in models of care combining elements of face-to-face and remote interaction; patient-centred and provider-support, and home and facility-based care. This accords with the articulated government position of support for greater involvement of patients in determining their care pathways(22) and that of shared decision making endorsed by the National Institute of Health and Care Excellence.(23) The accumulating body of evidence makes a powerful case for establishing the use of mifepristone at home with medical abortion via telemedicine as a permanent option in the repertoire of abortion provision.

## Data Availability

All data produced in the present study are available upon reasonable request to the authors

## ACKNOWLEDGMENTS

We are immensely grateful to all the women who dedicated their time and agreed to share with us their thoughts and experiences.

Thank you to the SACHA Study team as a whole for contributing ideas that were instrumental to this research: Annette Aronsson, Paula Baraitser, Caroline Free, Louise Keogh, Clare Murphy, Wendy Norman, Jill Shawe, Sally Sheldon, and Geoffrey Wong.

Thank you to the Advisory Group for supporting the development of this study.

